# Anti-vascular endothelial growth factor for diabetic macular edema: a Bayesian network analysis

**DOI:** 10.1101/2022.06.09.22276181

**Authors:** Xianhuai Wang, Xinyu Guo, Tianhui Li, Xincheng Sun

**Affiliations:** Department of Ophthalmology, The Affiliated Changzhou No.2 People’s Hospital of Ophthalmology, Changzhou, China; Department of Ophthalmology, Graduate School of Dalian Medical University, Dalian, China

**Keywords:** Anti-vascular endothelial growth factor, diabetic macular edema, Bayesian network meta-analysis, best corrected visual acuity, central macular thickness

## Abstract

**Aims:** Comparison of the efficacy of six Anti-vascular endothelial growth drugs in the treatment of diabetic macular edema

**Methods:** This network meta-analysis has been registered on the PROSPERO platform (CRD42022295684).A comprehensive search of eight databases without language restrictions.PubMed, EMBASE, Web of Science, Cochrane Library, CBM, CNKI, VIP, and Wanfang were used to search for randomized controlled trials on anti-vascular endothelial growth factor of diabetic macular edema, no language restrictions and publication time restrictions. According to the inclusion and exclusion criteria, literatures were screened, data were extracted and literature quality was evaluated, and the mean changes in best-corrected visual acuity (BCVA) and central macular thickness (CMT) were obtained. Using the Gemtc 1.0-1 package in R 4.1.2 to call the JAGS and the Markov Chain-Monte Card (MC-MC) method for Bayesian network meta-analysis. Risk of bias was assessed using RevMan 5.3. Stata 14.0 draws funnel plots to assess publication bias.

**RESULTS:** Sixteen studies including 3651 eyes were included, all with treatment follow-up over 6 months. The overall heterogeneity in the network analysis was low(I^²^=0), and there was no inconsistency. For the efficacy of reducing CMT, ranking according to the cumulative probability: Faricimab (0.9) > Brolucizumab (0.87) > Aflibercept (0.58) > Conbercept (0.37) > Ranibizumab (0.29)>Bevacizumab (0), but there was no statistically significant difference between Conbercept and Faclibercept, Aflibercept, and Buloxizumab. For the efficacy of improving BCVA, ranking according to the cumulative probability: Conbercept (0.87) > Faricimab (0.79) > Aflibercept (0.61) > Brolucizumab (0.51) > Ranibizumab (0.2)>Bevacizumab (0.02), but there was no statistically significant difference between Conbercept and Faclibercept, Aflibercept, and Brolucizumab.

## Introduction

Diabetic retinopathy (DR) is one of the most common and serious complications of diabetes mellitus (DM)[1], and diabetic macular edema (DME) is due to the damage of the retinal barrier caused by increased retinal vascular permeability in diabetic patients. This can lead to macular edema[2]. Macular edema can lead to significant loss of vision, and long-term edema can lead to irreversible damage to vision. Vision loss is also the most common symptom of diabetic macular edema. The current gold standard for the detection of diabetic macular edema is optical coherence tomography (OCT), which not only clearly shows macular thickening but also has the advantages of reproducibility and non-invasiveness[3]. With the continuous research and development of drugs, more and more randomized trials have shown that intravitreal injection of anti-vascular endothelial growth factor(VEGF) has largely replaced retinal laser photocoagulation[4]. Steroid injection is also a treatment for diabetic macular. The method of edema is suitable for the case of ineffective or resistant anti-vascular endothelial growth factor, and may lead to the early occurrence of cataract and the adverse consequences of ocular hypertension[5]. Therefore, the current safe and effective first-line treatment for diabetic macular edema is intravitreal injection of anti-vascular endothelial growth factor[6].

At present, anti-vascular endothelial growth factor mainly include Ranibizumab, Aflibercept, Conbercept, and Bevacizumab. These four drugs have been widely used in clinical practice, and two emerging drugs, Brolucizumab and Faricimab, were recently listed by the U.S. Food and Drug Administration (FDA). Ranibizumab and Aflibercept were approved by the FDA in 2006 and 2010, respectively. Conbercept, an anti-neovascular drug independently developed in China, was approved for marketing in 2014. Bevacizumab for the treatment of DME belongs to off-label use. Although it is questioned based on supervision and safety, it is still selected by some countries and regions due to its low cost[7]. Brolucizumab was approved for the treatment of wet age-related macular degeneration (AMD) in 2019, and the results of a phase III clinical trial for the treatment of DME have been published[8]. Faricimab has been approved by the FDA for the treatment of AMD and DME in 2022. The efficacy of these drugs varies, and there is a lack of evidence for some direct comparisons, making it difficult to assess efficacy comparisons between them. In order to meet the needs of clinical practice, this study used the method of network meta-analysis to explore the efficacy of different drugs. This study included a comparison of six anti-vascular endothelial growth factor, which was more comprehensive than similar studies, and excluded the effects of interventions such as steroid injection and laser photocoagulation on outcome indicators. Patients were followed up for 6 to 12 months in all included RCTs, ensuring the reliability of the results.

## 2. Methods

The study was conducted according to the guidelines of the Cochrane Multiple Intervention Methods Group,the study was a randomised controlled trial (RCT) involving DME patients requiring anti-VEGF therapy. The type of intervention was a direct comparison between different anti-VEGF drugs, and steroid comparison studies were excluded. With regard to drug dose and duration, the included trials were all commonly used doses and regimens of drugs. Outcomes were the mean change from baseline to the end of follow-up in best-corrected visual acuity (BCVA) and the mean change from baseline to the end of follow-up in central macular thickness (CMT) measured using optical coherence tomography (OCT). The different measurement units of visual acuity are converted into a unified ETDRS letter score by formulas and then statistically calculated[9]. In case of different follow-up time, choose the follow-up time closest to 12 months.

### Literature search strategy

Comprehensive search of PubMed, EMBASE, Web of Science, Cochrane Library, CBM, CNKI, VIP and Wanfang databases. To identify relevant studies published prior to February 2022, without language and time of publication restrictions, using search terms (diabetic macular edema) and (ranibizumab or bevacizumab or aflibercept or conbercept or brolucizumab or faricimab) and (randomized controlled trial or RCT or random). All retrieved literature was reviewed and manual searches were performed to ensure completeness of references.

### Eligibility Criteria

The included literature must meet the following criteria: 1. The research subjects meet the diagnostic criteria for type I or II diabetes mellitus complicated with diabetic macular edema; 2. The intervention measures can only be six anti-VEGF drugs, excluding combination therapy and steroid therapy. 3. The outcome indicators of the study included BCVA and CMT and the follow-up time was more than 6 months. 4. The study design was a randomized controlled trial, and retrospective studies were excluded. 6. Some literatures with incomplete data, baseline data and unclear outcome indicators were excluded.

### Data extraction

Two reviewers (XH W,XY G) independently selected the included studies and extracted data. For the retrieved literature, use NoteExpress 3.5 to remove duplicate literature. After the initial study selection was completed, the studies to be included were compared.Consensus was reached after discussions. Two reviewers independently extracted follow-up time, interventions, mean (MD) and standard deviation (SD) of change in outcome measures, sex and age of patients in each study using Excel2019. Each reviewer independently read the entire article and finally compared the extracted data. In the case of some studies that did not provide the standard deviation directly, and the standard deviation could not be obtained from the authors, statistical formulas were used to calculate or use the baseline standard deviation. If it was not available, it could not be included in the analysis.

### Risk of Bias Within Individual Studies

Three reviewers (XH W, XY G, TH L) independently reviewed the included studies, using the Cochrane Risk of Bias Tool to assess the risk of bias, and assessed in the following aspects: 1) Whether random assignment was mentioned, and the manner of random assignment Whether it is correct; 2) Whether the concealed grouping is mentioned, and whether the concealed grouping method is correct; 3) Whether the researchers and subjects are blinded, and whether the blinding is correct; 4) The integrity of the result data; 5) Selective reporting of results; 6) Other biases, such as those caused by receiving funds, deceptive behavior, etc. For each aspect the evaluation was set to low risk, high risk and unclear. Risk of bias was assessed using RevMan 5.3 and a risk of bias map was drawn.

### Sensitivity and Subgroup Analysis

In order to prove the stability of the results, sensitivity analysis can be performed on the efficacy evaluation according to the divided follow-up time to observe whether there is consistency among the groups. In DRCRnet2015[10], the authors mentioned that the patient’s treatment benefit depends on baseline best visual acuity, or baseline CMT. Subgroup analyses can be discussed based on patients’ baseline BCVA or CMT.

### Statistical methods

Stata 14.0 was used to draw a network graph to present the relationship between direct and indirect comparisons of drugs among different studies, and an inverted funnel plot was used to evaluate whether the interventions had small sample effects or publication bias. Using the Gemtc 1.0-1 package in R 4.1.2 to call JAGS software and the Markov Chain-Monte Card (MC-MC) method to conduct Bayesian network meta-analysis. And use the ggplot2 package to draw the cumulative probability curve and the reference graph in the mixed case of the two outcome indicators. Four Markov chains were used for the simulation, and the number of iterations was set to 50,000 times (the first 20,000 times were used for annealing to remove the effect of the initial value, and the last 30,000 times were used for sampling). The iterative convergence was judged by calculating the potential scale reduction factor (PSRF) and the Gelman Rubin Brooks diagnostic method. The median value of the reduction factor and 97.5% tended to 1 and reached stability after n iterations of calculation, and the PSRF was limited to 1 to 1.05. Satisfactory convergence is achieved. Global inconsistency is judged by using the value of the deviation information criterion (DIC) between consistent and inconsistent models. If the difference in DIC is within 5, the data are generally considered to be consistent, and a fixed-effects model is used; if the difference is greater than 5, the data is considered inconsistent and a random-effects model is used. The local consistency of each closed loop is tested by the node splitting method. If the p-value of the local consistency test is greater than 0.05, it indicates that the consistency between the direct comparison and the indirect comparison is low, and the consistency model is used for analysis. Otherwise, the closed loop is considered to have obvious inconsistency.

## Results

### Essential characteristics of the included studies

Fig 1 shows the process of article selection. A total of 2076 records were obtained from the database search. After excluding duplicate literature, further selection was made by reading the full text, and those that did not meet the intervention measures, did not meet the study design, and had insufficient follow-up time were excluded. Finally, sixteen studies were included.

**Fig 1.**
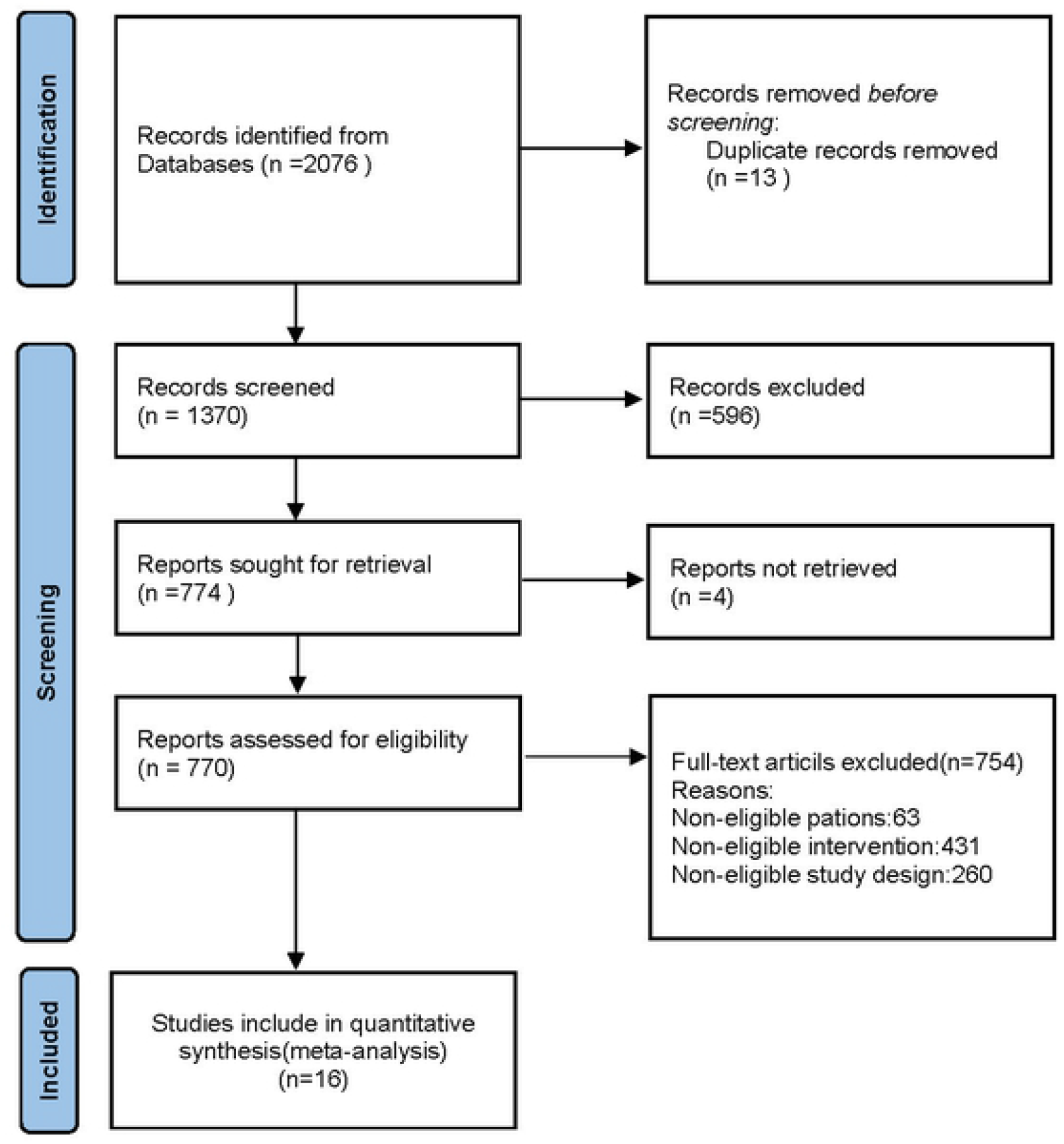
Flow chart of the selection process for this network meta-analysis.

### Literature Features and Network Evidence Map

A total of 16 studies were included in the network analysis. The basic characteristics o f each study’s intervention measures and the number of participants can be found in Table 1. Fifteen studies were two-arm studies, and one study was a three-arm study. Six -drug regimens: aflibercept, bevacizumab, ranibizumab, conbercept, buloxacin, and farisimumab, a total of 3651 eyes were randomly assigned to anti-vegf therapy, eigh stuies(Khaled2020[11],Qinsy2020[12],Vader2020[13],Shenhm2021[14],DuanM M2016[15],Jayashree2019[16]) were followed up for six months, and the rest of the stuies(Ekinci2014[17],Alexandros2020[18],Fouda2017[19],Nepomuc2013[20],Wiley20 16[21],YOSEMITE2022[22]) were followed up for more than six months months, and the time closest to twelve months was taken as the end point of follow-up. In Qinsy20 20[12], the results in the text are that the BCVA of the experimental group and the control group after treatment is lower than that before treatment, so the logMAR statistical visual acuity index used by the author is considered, but the mean difference after conversion to ETDRS calculation reaches 59 letters, which is very different from other studies, and the mean of the baseline logMAR visual acuity of the observation group i s 1.62, which is already greater than the maximum logMAR visual acuity of 1.6. A detailed analysis of the article and attempts to contact the authors did not lead to reasonable conclusions, so the BCVA change from this study was not included in the analysis.

Finally, A network graph of the two outcome indicators was constructed. Fig2. Network graph summary: dots indicate interventions (A: Aflibercept B: Bevacizumab R: Ranibizumab C: Conbercept L: Brolucizumab F: Faricimab), The larger the dots, the more patients receiving the drug; the straight line indicates that there is a direct comparison between the two drugs. A total of 14 studies involved ranibizumab in this analysis, the most of any drug. The thicker the line, the greater the number of studies in which the two drugs were directly compared, with the most comparisons between Ranibizumab and Bevacizumab at six.

**Fig 2A.**
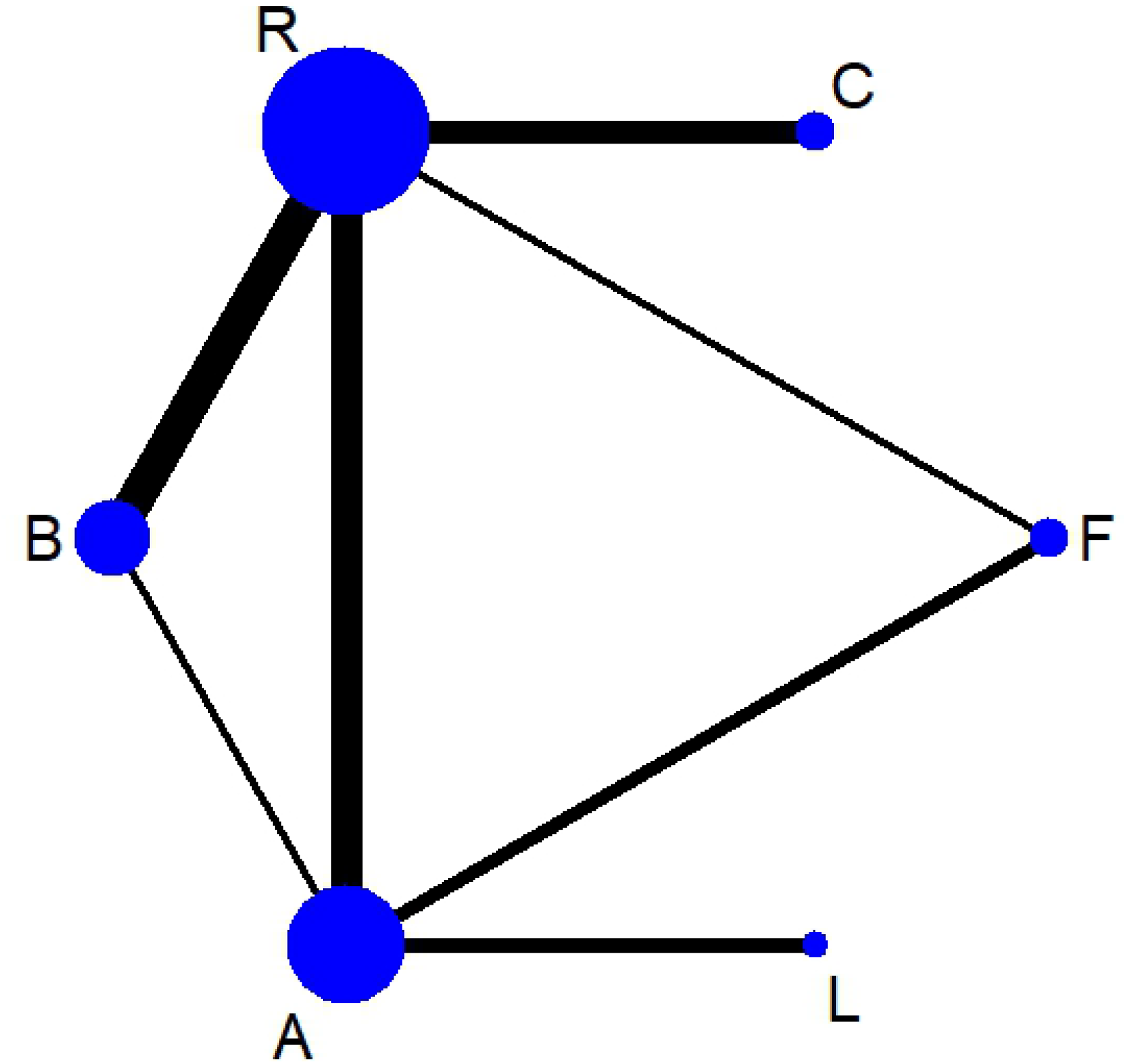
Network plot of CMT. A: Aflibercept B: Bevacizumab R: Ranibizumab C: Conbercept L: Brolucizumab F: Faricimab

**Fig 2B.**
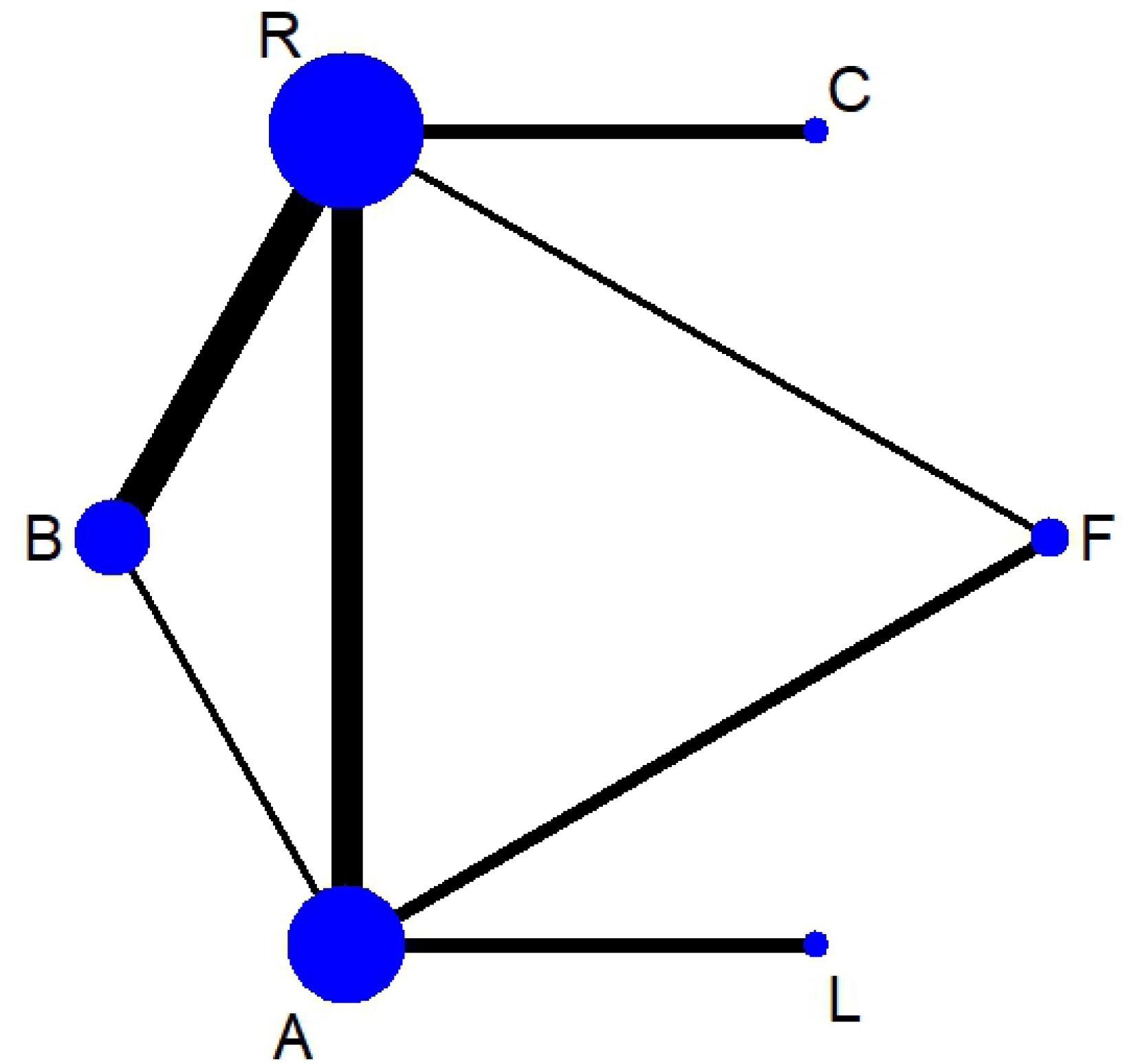
Network plot of BCVA. A: Aflibercept B: Bevacizumab R: Ranibizumab C: Conbercept L: Brolucizumab F: Faricimab

### Quality assessment and risk of bias assessment

The study quality assessment table included in this network meta-analysis is shown in the S1 and S2 File, and assessed in terms of random sequence generation, allocation concealment, blinding implementation, data integrity, and selection reporting. Most were rated as low risk of bias and unclear risk of bias. To analyze the publication bias in the studies, the horizontal axis represents the effect size of each study, and the vertical axis represents the standard error of each study to draw a corrected funnel plot(Fig 3). The overall funnel plot was well symmetric, indicating little publication bias.

**Fig 3A.**
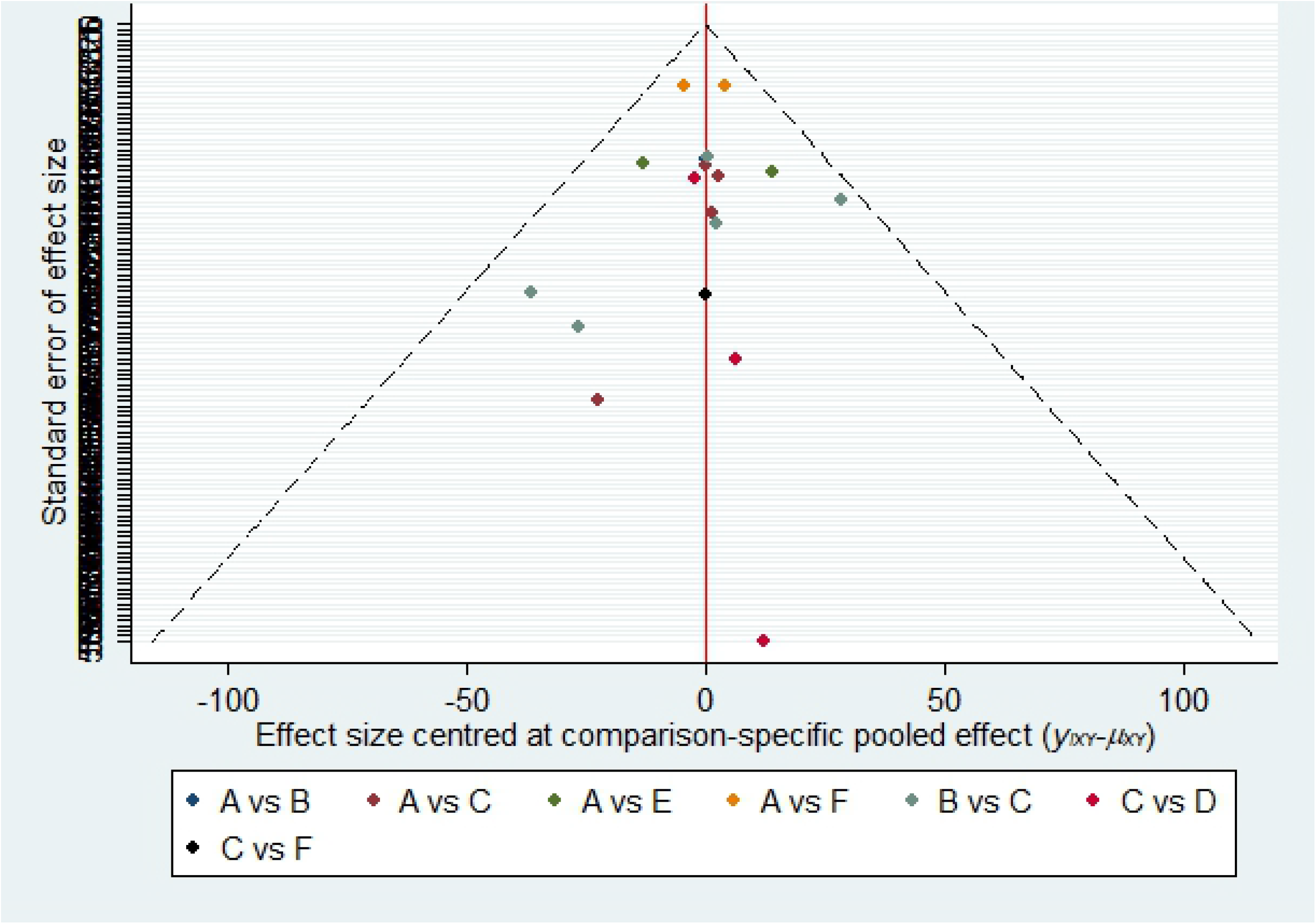
Risk of bias funnel plot for CMT. The horizontal axis represents the effect size of each study, and the vertical axis represents the standard error of each study.

**Fig 3B.**
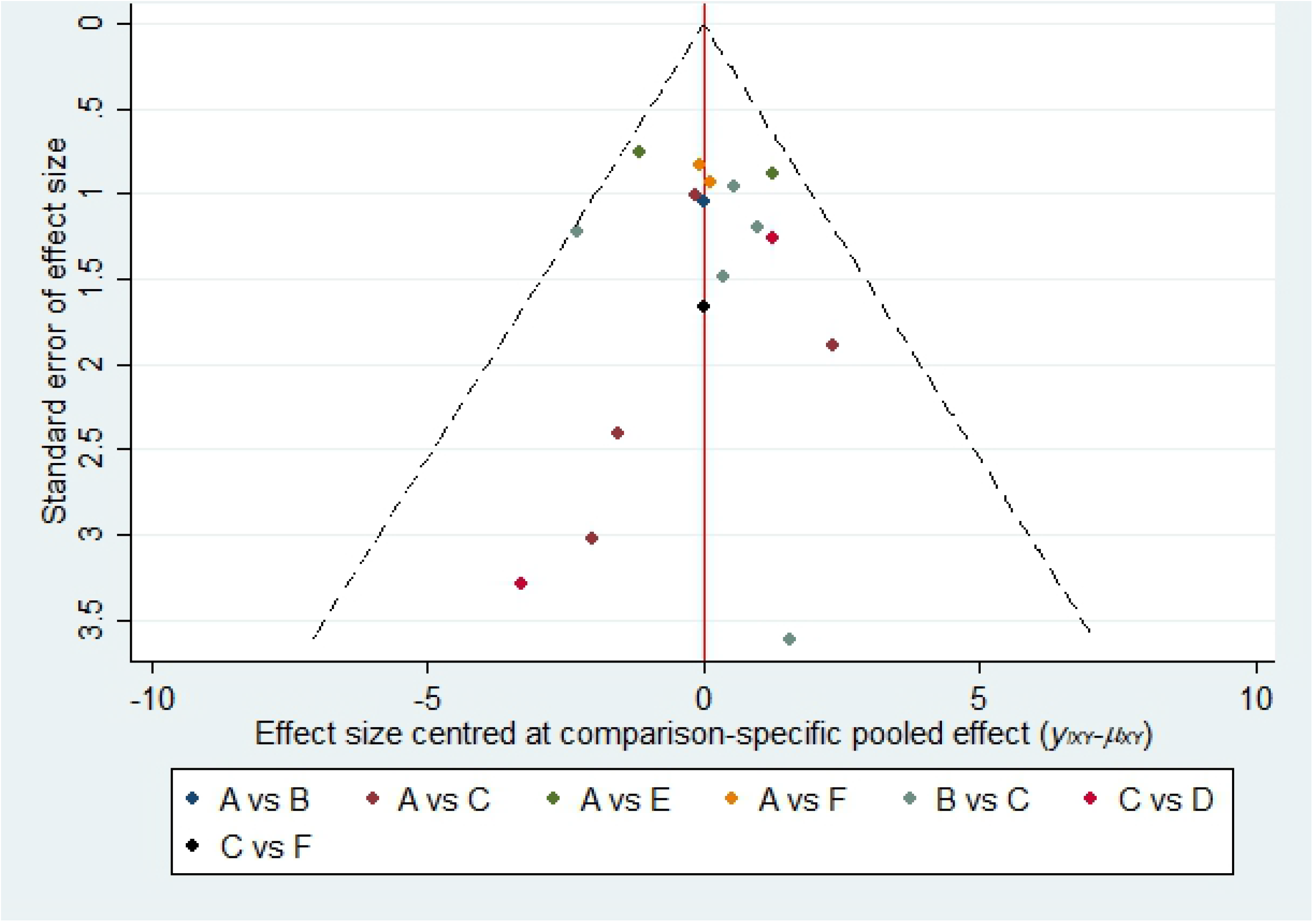
Risk of bias funnel plot for BCVA. The horizontal axis represents the effect size of each study, and the vertical axis represents the standard error of each study.

### In reducing central retinal thickness (CMT)

All anti-vegf drugs have a reduction in central macular thickness, Figs 4 shows the ranking cumulative probability of CMT reduction for all drugs, and the area under the curve represents the cumulative probability of ranking, specifically: Farecimab (0.9) > Brolucizumab (0.87) > Aflibercept (0.58) > Conbercept (0.37) > Ranibizumab (0.29) > Bevacizumab (0). However, according to the pairwise comparison between different drugs in the league table(Table2) (upper right corner), there is no statistically significant difference between Farecizumab and Conbercept and Brolucizumab. There was also no statistically significant difference between buloxetine and aflibercept and conbercept. For details, see Table 2, which is presented in the form of MD 95%Crl. The results are interpreted as comparing the effect size with the drug corresponding to each row, and the green represents no statistically significant difference.

**Fig 4.**
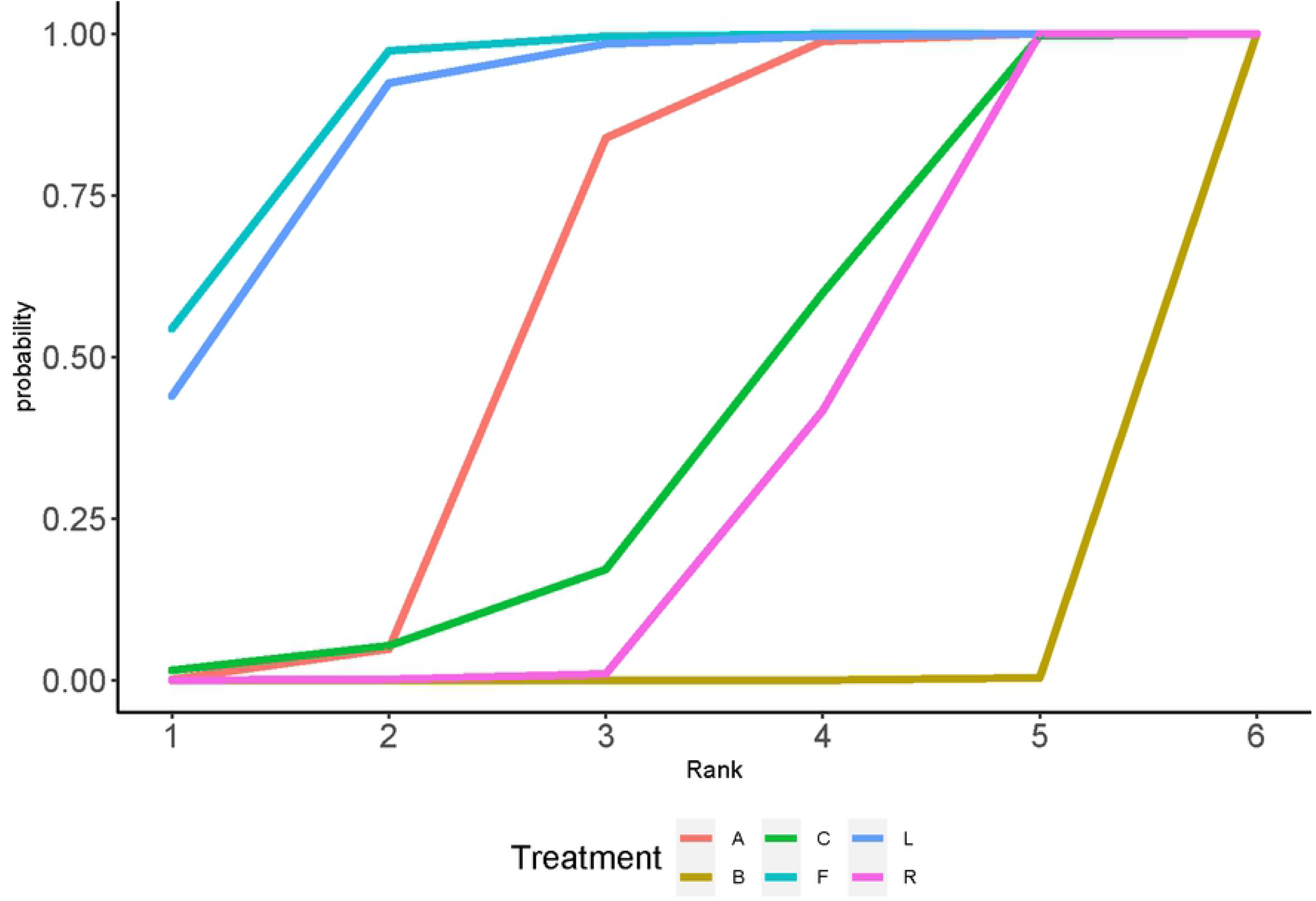
Cumulative probability plot of CMT. The area under the curve represents the cumulative probability of ranking.

**Table 1.**
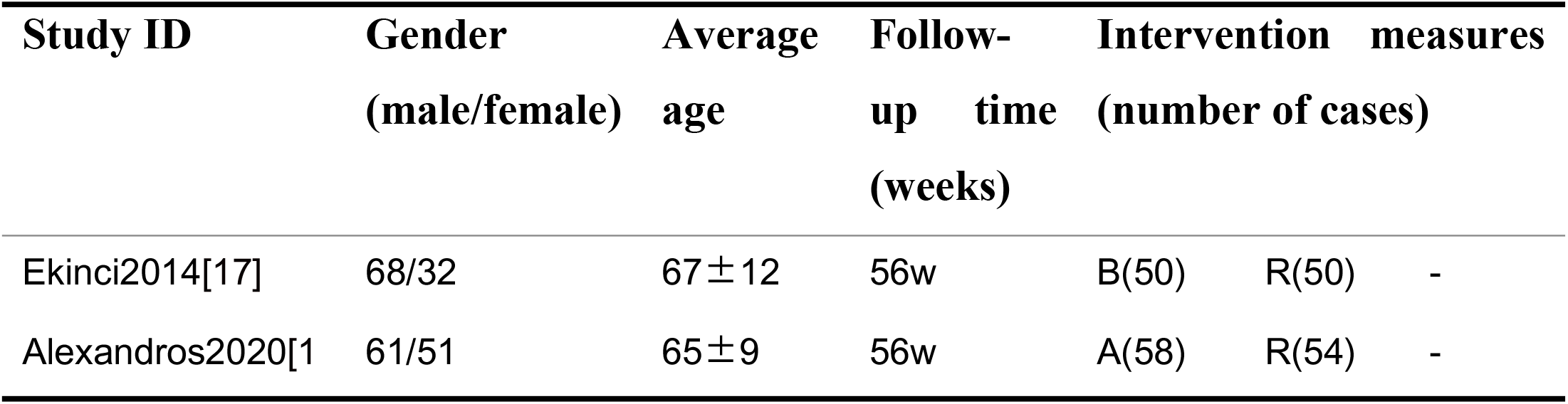
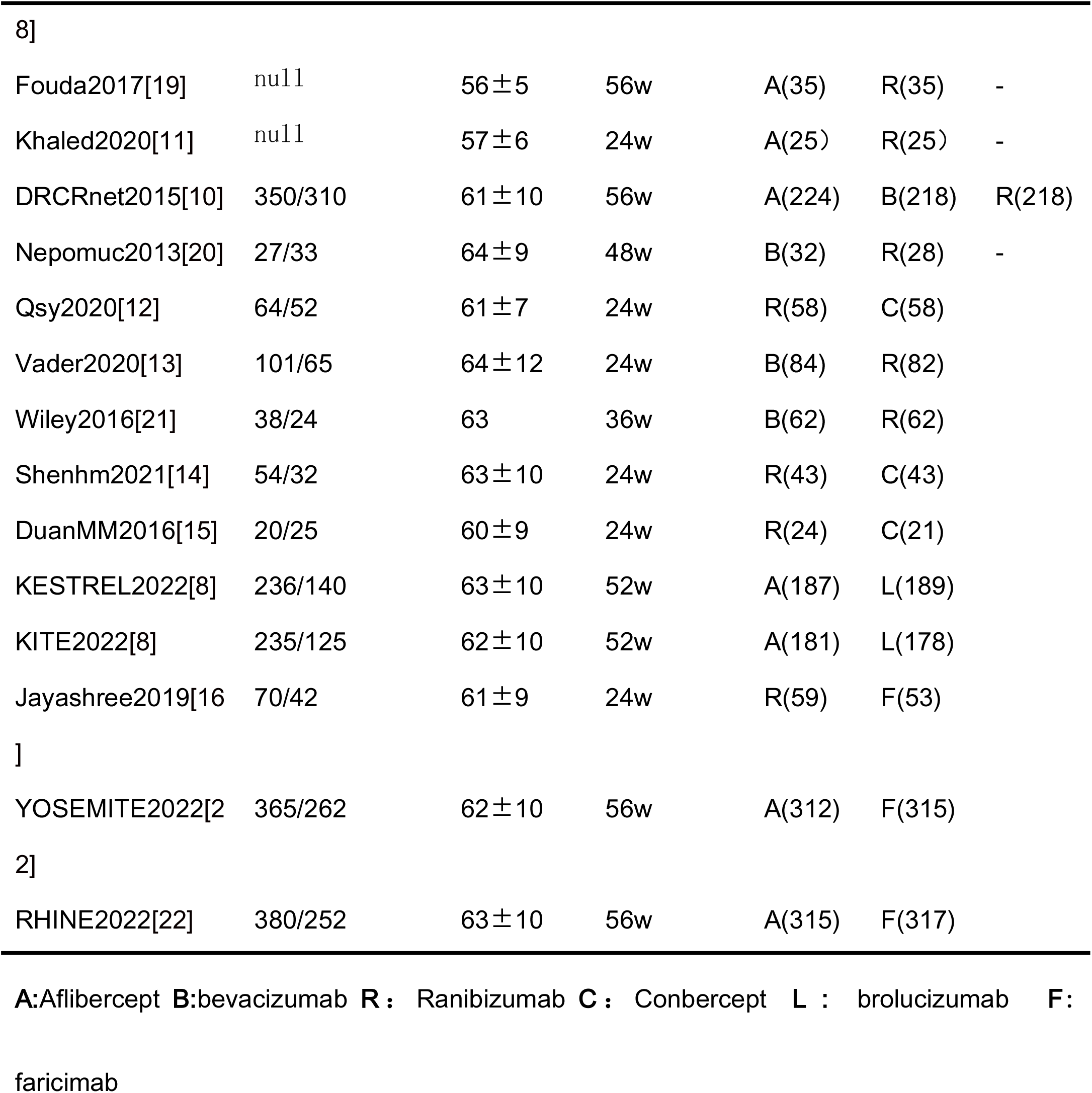
Characteristics of the included studies of the network meta-analysis.

**Table 2.** Pairwise comparisons between different drugs.

|  | A | B | C | F | L | R |
| --- | --- | --- | --- | --- | --- | --- |
| A | A | -66.63 (-88.5, -42.99) | -16.17(-50.5,18.51) | 20.9 (4.17, 34.93) | 18.95(-4.06, 42.03) | -19.92(-37.65, -1.52) |
| B | 3.2 (1.06, 5.4) | B | 50.34(15.19,84.28) | 87.39(59.3,112.47) | 85.62(52.38,117.14) | 46.59 (28.51, 63.96) |
| C | -1.65 (-5.02, 2) | -4.86 (-8.09, -1.34) | C | 36.9 (-0.84, 73.1) | 35.29 (-6.44, 76.5) | -3.57(-33.28, 25.47) |
| F | -0.72(-2.61,1.07) | -3.94 (-6.63, -1.35) | 0.93 (-3.09, 4.53) | F | -1.75(-28.63,27.03) | -40.55(-61.99,17.58) |
| L | 0.41(-1.65,2.34) | -2.8 (-5.83, 0.07) | 2.07 (-2.2, 5.9) | 1.13(-1.58, 3.81) | L | -38.89(-68.22,-9.73) |
| R | 2.26 (0.44, 4.1) | -0.95 (-2.51, 0.59) | 3.91 (0.75, 6.77) | 2.98 (0.72, 5.33) | 1.86 (-0.77, 4.6) | R |

### In Improving best corrected visual acuity (BCVA)

All anti-vegf drugs have improved visual acuity, and the area under the curve in Fig 5 shows the cumulative probability of the ranking of all drugs on visual improvement, specifically: Conbercept (0.87)>Farisimumab (0.79)> Aflibercept (0.61) > Brolucizumab (0.51) > Ranibizumab (0.2) > Bevacizumab (0.02). However, according to the league table, there was no statistically significant difference between Conbercept and faricilimab, aflibercept, and brolucizumab. The specific information is shown in the lower left corner of Table 2, which is presented in the form of MD and 95%Crl. The results are interpreted as the comparison effect size of the drug corresponding to each row. Green indicates that the difference is not statistically significant.

**Fig 5.**
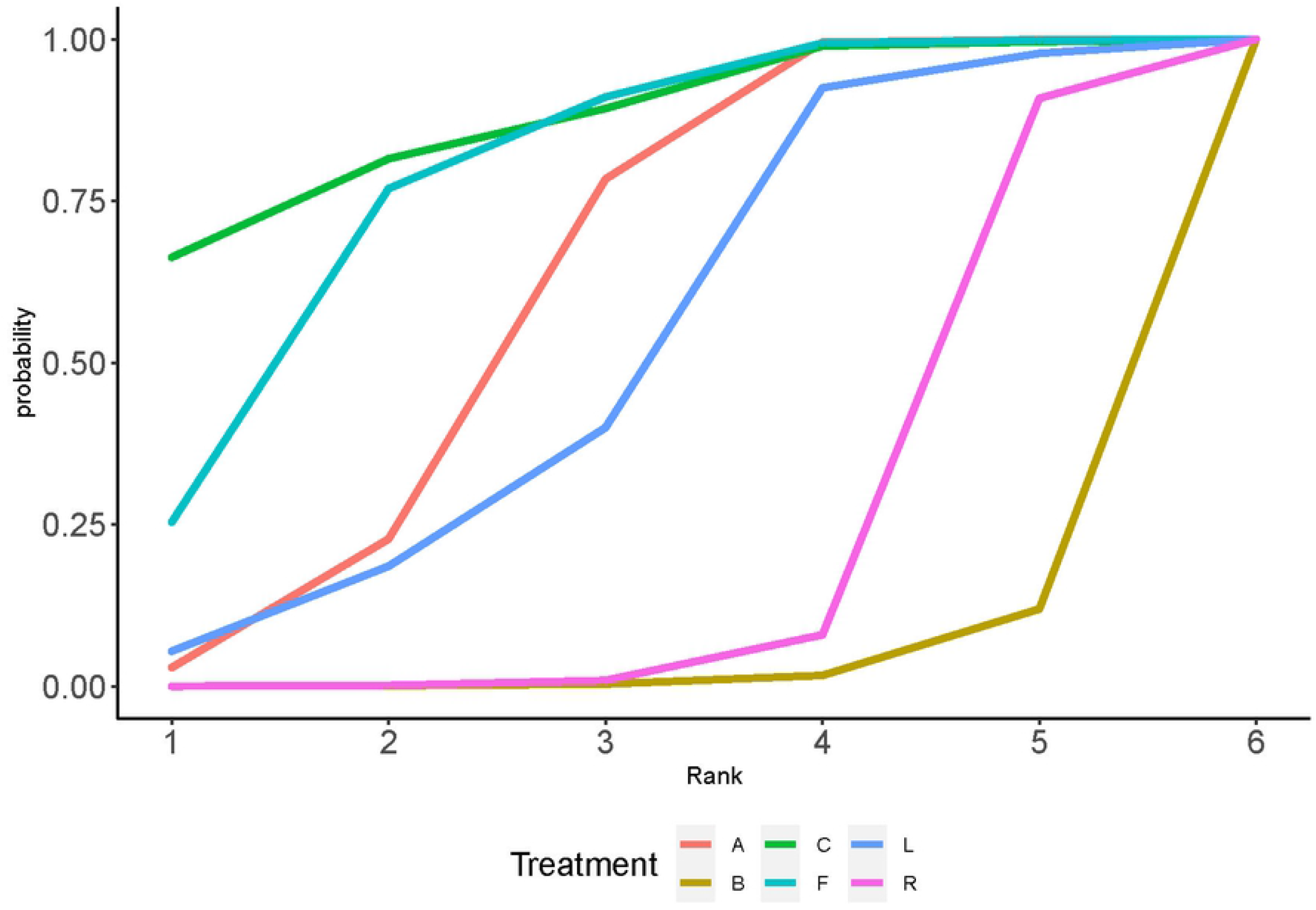
Cumulative probability plot of BCVA. The area under the curve represents the cumulative probability of ranking.

### Combining two outcome measures

When measuring two outcome indicators at the same time, Bevacizumab was used as the reference, the relative change in CMT was used as the abscissa, and the BCVA relative to this variable was used as the ordinate, as shown in Fig 6. Faricimab have certain advantages in both BCVA and CMT improvement.

**Fig 6.**
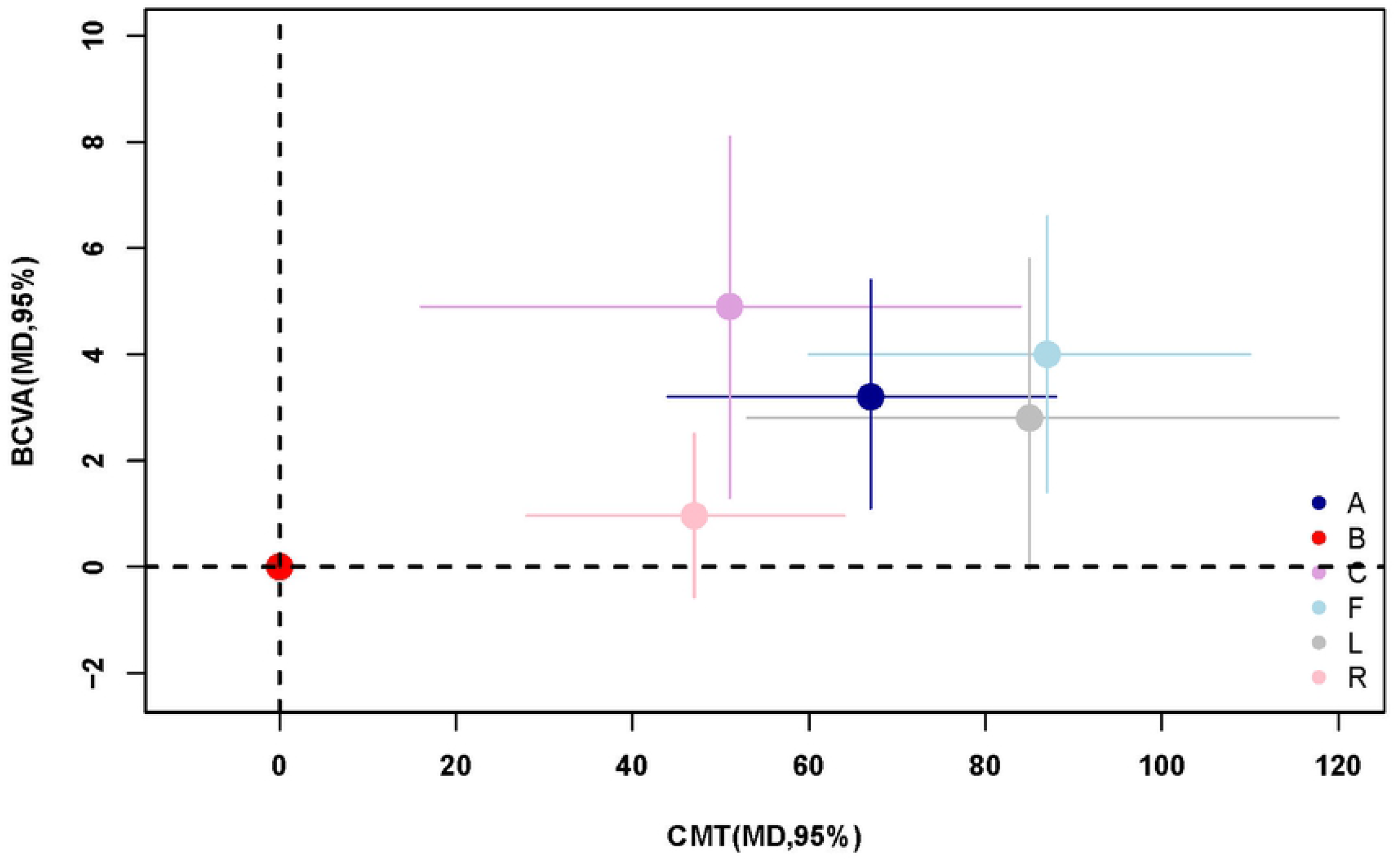
Mixed Efficacy Plot of CMT and BCVA. The relative change in CMT was used as the abscissa, and the BCVA relative to this variable was used as the ordinate.

### Heterogeneity analysis

The two outcome indicators were tested for heterogeneity, and there was no obvious heterogeneity in the overall population. For details, see S3 and S4 File. The average BCVA changes: the overall heterogeneity I^²^=0%, and the difference between some studies. The higher quality is due to the direct comparison of the results, as in the comparison between Brolucizumab and Aflibercept, there were only two direct comparison studies between them, and both were Brolucizumab Phase III clinical trials are being conducted concurrently. The final conclusion from the results of both studies was that Brolucizumab and Aflibercept had comparable BCVA improvement in patients with DME, which is consistent with our net comparison results. In the mean CMT change: the overall heterogeneity I^²^=0%, the overall heterogeneity is also good.

### Local inconsistency analysis

The local inconsistency analysis was performed using the node separation method. In the two outcome indicators, the network evidence map showed that there were two closed-loop studies, and the two analysis results showed that the P-value between all studies was greater than 0.05, indicating that the results of direct comparison and indirect comparison between all drugs in the closed loop are consistent, and there is no inconsistency, see the S5 and S6 File for details.

### Sensitivity

In order to keep the results stable, we have excluded studies with a follow-up period of less than six months when we included the analysis. We tried to divide the analysis between six months and twelve months to see whether the follow-up period affected the overall results, However, it was found that the number of studies after division was insufficient for a network meta-analysis. Sensitivity analysis was performed in part of the meta-analysis on anti-vegf treatment of DME, and the sensitivity analysis in Xuexue Zhang2021[23] showed that different follow-up times were not found to significantly affect the results of BCVA and CMT. Similarly WeiShai Liu2019[24], the sensitivity analysis reached the same conclusion.

## Discussion

This is the first comparative study to evaluate and compare the head-to-head effectiveness of six anti-VEGF drugs, and compared to previous meta-analyses, this analysis not only includes four drugs that have been used on a large scale in clinical practice (Conbercept, Aflibercept, Ranibizumab, and Bevacizumab) also include drugs that have undergone phase III clinical trials but have not yet entered clinical practice on a large scale (Brolucizumab, Faricimab). In addition, the interference of steroid drug injection and laser photocoagulation therapy is excluded, because these two treatment measures are not the first choice for the treatment of DME, and are only used as supplementary therapy for anti-vegf[25]. According to the statistical results, Faricimab is a relatively better choice in changing CMT. As the first dual-pathway-specific antibody designed for ophthalmology, fariximab is angiopoietin-2 (Ang -2) and vascular endothelial growth factor-A (VEGF-A). These two receptors can increase vascular permeability and promote the formation of macular edema[26]. Conbercept was a relatively better choice for BCVA,but did not differ significantly from Faricilumab. Conbercept is a new type of anti-VEGF drug developed and produced in China, which has not yet been widely used in the world. Meanwhile, the large-scale clinical trials of Conbercept are not yet completed. This result still needs more research to evidence, but some advantages of Conbercept have also been shown in some retrospective clinical studies[27]. For several other drugs that have been used on a large scale in clinical practice, all efficacy results of Aflibercept are better than Ranibizumab and Bevacizumab, which is basically consistent with the findings of DRCRnet 2015[10]. The inclusion of multiple studies and based on indirect comparisons in Bayesian network meta-analyses reinforced the conclusions of several large-scale clinical head-to-head direct comparisons because there were no apparent inconsistencies. Further showing the advantages of Faricimab.

### Limitations

This study only analyzed the efficacy of the drug, not the safety of the drug, because some studies did not report safety. However, based on the retrospective report ChengY2021[28] and the safety of intravitreal drug injection in clinical trials, there was no significant difference[25]. In addition, the existing literature is not comprehensive enough, and the quality of the literature is also uneven, limited by differences in medical levels in different places, and there are differences in the measurement of some outcome indicators, which may cause differences in the study results, although in the end overall heterogeneity does not high. This network meta-analysis did not completely unify the follow-up time, and only relatively selected studies with a follow-up time of more than six months in order to eliminate the difference in the results due to the follow-up time, but it may still have an impact on the results. As for the efficacy evaluation of the drugs after one year, only the Protocol T trials of Aflibercept, Ranibizumab and Bevacizumab are relatively complete. At two years, all three anti-VEGF drugs showed BCVA from baseline to two years improvement in the second year, and fewer injections in the second year. Vision outcomes were similar in eyes with better baseline BCVA. In poorer baseline visual acuity, Aflibercept was associated with better 2-year BCVA outcomes compared with Bevacizumab, but the advantage of Aflibercept over Ranibizumab was no longer established[29]. At five years, although CMT was similar at two and five years, the mean BCVA worsened during this period, and the specific treatment regimen could not be determined, and further investigation is needed[30]. Subgroup analyses were not performed by baseline BCVA, baseline CMT, or by type of diabetes because the number of head-to-head clinical trials between drugs was not sufficient for subgroup analyses, and details of type of diabetes were not available in every literature. Therefore, the impact of these differences on the results could not be assessed. Additional high-quality randomized controlled trials are needed to fully and accurately evaluate these drugs.

## Conclusions

This network meta-analysis analyzed the efficacy of different anti-vegf drugs in the treatment of diabetic macular edema. In general, Faricimab was relatively more effective in improving CMT and BCVA. This conclusion may provide help for clinicians and adjust the treatment plan accordingly, but it still needs a lot of clinical practice to confirm.

### Registration and protocol

This network meta-analysis has been registered on the prospero platform, the registration number is: CRD42022295684,the registration title is: Four anti-vascular endothelial growth factor for diabetic macular edema:A Bayesian Network Analysis, any information can be reviewed on the prospero platform, after the analysis found two new anti-VEGF drugs, changed again Retrieve strategies and analyze them.

## Data Availability

All relevant data are within the manuscript and its Supporting Information files.

## Funding

This work is supported by Changzhou Municipal Health Commission Major Project(ZD201712).

## Authors’ contributions

Conception and design: XH W and XC S; Collection and assembly of data: XH W and XY G; Data analysis and interpretation: All authors; Manuscript writing: XH W; Final approval of manuscript: All authors; Accountable for all aspects of the work: All authors.

### Availability of data and materials

The authors confirm that the data supporting the findings of this study are available within the article.

Code availability

Not applicable.

### conflict of interest

The authors declare that the research was conducted in the absence of any business or financial relationships that could be construed as potential conflicts of interest

## Abbreviations

BCVA: Best corrected visual acuity
CMT: central macular thickness
DR: Diabetic retinopathy
DM: diabetes mellitus
DME: diabetic macular edema
VEGF: vascular endothelial growth factor
A: Aflibercept
B: bevacizumab
R: Ranibizumab
C: Conbercept
L: brolucizumab
F: faricimab

## Supporting information

**S1** File. Risk of bias graph.

**S2** File. Risk of bias summary.

**S3** File. Heterogeneity test for CMT.

**S4** File. Heterogeneity test for BCVA.

**S5** File. Local heterogeneity of CMT.

**S6** File. Local heterogeneity of BCVA.

## Notes

### Competing Interest Statement

The authors have declared no competing interest.

### Funding Statement

The funders had no role in study design, data collection and analysis, decision to publish, or preparation of the manuscript.

